# Swaasa an AI-Based Novel Technique to Assess Lung Function in Yoga Practitioners in Comparison to Non-Yoga Practitioners: A Cross-Sectional study

**DOI:** 10.64898/2026.11.05.26359313

**Authors:** Sobika Rao, Rameswar Pal, Amisha Rawat, Parul Chaudhury, Ishwara Nayarkere Acharya, Vikram Singh, Kashinath Samagandi, Shubha Deepti Palreddy

## Abstract

**Background:** Lung function is a key indicator of respiratory health and is known to be influenced by lifestyle practices such as yoga. Recent advances in artificial intelligence (AI) have enabled non-invasive, scalable, and real-time assessment of physiological parameters. Swaasa is an AI-based novel technique designed to assess lung function through cough analysis, offering a potential alternative to conventional spirometry.

**Objective:** The present study aimed to evaluate lung function using Swaasa among yoga practitioners and compare the findings with age- and sex-matched non-yoga practitioners in a cross-sectional design.

**Methods:** A cross-sectional comparative study was conducted on adult participants divided into two groups: regular yoga practitioners (≥1 year of practice) and non-yoga practitioners. Lung function parameters, including Forced Vital Capacity (FVC), Forced Expiratory Volume in one second (FEV1), and cough pattern indices, were assessed using the Swaasa AI-based system.

**Results:** Yoga practitioners demonstrated significantly higher AI-derived lung function indices, including FVC and FEV1, compared to non-yoga practitioners (p < 0.05). Improved respiratory health and reduced respiratory variability were also observed in the yoga group, indicating enhanced respiratory efficiency.

**Conclusion:** The findings suggest that regular yoga practice is associated with better lung function as assessed by Swaasa.

## Introduction

Respiratory health is a basis of preventive and rehabilitative medicine, particularly in the context of chronic respiratory diseases, post-infectious sequelae, and lifestyle-related pulmonary dysfunction. Also, respiratory disorders contribute substantially to global morbidity and mortality, placing a significant burden on health care systems worldwide. [1]

Spirometry, a routinely performed pulmonary function test, as the name suggests, is a measurement of the amount of air one breathes. It includes measurements of both the volume and the rate of air that can be inhaled and exhaled as a function of time and enables classification of respiratory disorders into obstructive, restrictive, or mixed patterns in accordance with American Thoracic Society and European Respiratory Society (ERS) guidelines. [2]

Recent advancements in the field of artificial intelligence (AI) and machine learning have led to the emergence of digital biomarkers for the detection of disorders of the respiratory system. Acoustic features of cough sounds have been shown to correlate with specific cough pathologies. [3,4] Swaasa® AI-powered respiratory assessment platform developed by Salcit Technologies Pvt. Ltd., Hyderabad, is a point-of-care technology to assess a user’s respiratory condition. It uses cough analysis as an additional factor in risk assessment. Using proprietary machine learning and AI algorithms, the platform analyzes cough acoustics in combination with selected symptom inputs derived from the St George’s Respiratory Questionnaire (SGRQ). Subject to coughing in the mobile, within approximately 10 seconds, the platform identifies underlying respiratory conditions, the pattern of cough (obstructive, restrictive, or normal), and its severity. The Swaasa AI platform also provides the functionality of spirometry without the constraints of time, space, specialized equipment, and trained professionals.

Yoga is a 5,000-year-old tradition from India that integrates physical, mental, and spiritual pursuits to promote physical, mental, and physiological balance.[5] Pranayama practices influence autonomic nervous system balance, respiratory muscle strength, lung compliance, and gaseous exchange efficiency. [6,7]

As per the WHO, chronic obstructive pulmonary disease (COPD) is a major global health concern and is projected to become the third leading cause of death worldwide by 2030. [8] The Global Initiative for Chronic Obstructive Lung Disease (GOLD) emphasizes the role of non-pharmacological interventions, including reduction in symptoms, complications, exacerbations, improved exercise tolerance, improved health status, and reduced mortality. [9] Evidence suggests that yoga-based breathing interventions improve diffusion capacity, reduce associated stress and anxiety, and help improve quality of life in such patients. [10,11]

Asthma affects approximately 300 million individuals globally and is a significant cause of morbidity, particularly in low- and middle-income countries. [12] Clinical studies report significant improvements in Peak Expiratory Flow Rate (PEFR), Forced Expiratory Volume 1 (FEV1), Forced Vital Capacity (FVC), Vital Capacity (VC), inflammatory markers, and an absolute eosinophil count following breathing interventions. [13,14]

The post-Covid respiratory era has emerged as a major clinical challenge. The patients frequently experience persistent dyspnea, decreased lung function, persistent fatigue, and respiratory symptoms, impaired functional capacity, and diminished quality of life for months after acute infection. Radiological abnormalities and pulmonary function impairments have been documented up to six months after hospital discharge, even in individuals with non-severe disease. [15]

Coughing is a powerful and effective defense mechanism to protect the lungs. Leveraging cough as a digital biomarker, the Swaasa AI platform aims to bridge gaps in accessibility, affordability, and scalability, offering location-independent, equipment-free respiratory health assessment. Various validation studies have been done reporting the platform’s high sensitivity (95.45%) for COVID-19 screening 20, strong diagnostic accuracy (98.82%) for pulmonary tuberculosis screening, and highlighting the platform’s feasibility for large-scale population screening.[16,17]

When traditional yoga adopts new technology like AI, practitioners are in better control and can monitor and assess their breathing and lung condition. Nonetheless, there is evidence of growing yoga suggesting the benefits of yoga on respiratory health, but a comprehensive and controlled examination is necessary to elucidate the extent to which yoga can influence respiratory parameters. Therefore, integrating AI-enabled respiratory assessment with yoga practices presents a novel, evidence-based approach to assessing the potential benefits of yoga on respiratory health. Also, despite growing evidence supporting both yoga-based respiratory interventions and AI-driven cough analysis independently, studies evaluating their combined application remain limited.

Therefore, the present study aims to assess the lung health of yoga practitioners and non-yoga (sedentary) practitioners using the Swaasa AI platform and subsequently conduct a large scale randomized controlled trial to evaluate the effectiveness of pranayama and selected yoga practices on lung health indices across different respiratory conditions.

### Methodology

#### Ethical considerations

The study was conducted at the Morarji Desai National Institute of Yoga (MDNIY), Ministry of Ayush, Government of India, New Delhi, India. The study protocol was submitted to the Institutional Faculty-Cum-Research Committee (IFCRC) of MDNIY and was reviewed and discussed by the committee on 15 March 2024. Following the review and discussion, the committee granted approval for conducting the study. The approval was obtained before the commencement of the study. Written informed consent was obtained from all participants before their participation in the study.

#### Study Setting and Participants

The study was conducted in Morarji Desai National Institute of Yoga (MDNIY), Ministry of Ayush, Government of India, New Delhi. An announcement was made about the ongoing project, and voluntary participation was invited from the staff and students of MDNIY only. Healthy subjects from both genders, aged between 21-40 years, were included. The participants were screened for seasonal allergies and any acute episode of respiratory disorder. Participants who were taking any medication for a respiratory condition during the course of data collection were excluded. Pregnant women were also excluded from the study. Based on voluntary participation, the subjects were divided into two groups: Group A—who were yoga practitioners—and Group B, who were non-yoga practitioners (sedentary people).

#### Parameters

Swaasa AI platform was used to analyze cough signals along with the subset of symptoms from SGRQ (St. George’s Respiratory Questionnaire). A demographic form was also given to the subjects. Habits (smoking/alcohol/drugs) and history of covid-19 exposure were taken from the demographic form. Outcome measures: Risk, Pattern, Pattern severity, Lung Health Index, Asthma likely, Asthma Risk, COPD likely, COPD Risk, FEV1/FVC ratio, FEV1, FVC, and symptoms such as mucus/sputum, cough during night, difficulty in breathing, wheezing, frequent cough, family history of asthma, and exposure to any smoke were recorded using the Swaasa AI cough sound analyzer and questionnaire.

#### Statistical Analysis

The data was analyzed by calculating the score in terms of frequency, percentage mean, deviation, chi-square and unpaired T-test. Analysis and interpretation of data was done according to the objectives using descriptive and inferential statistics. The level of significance chosen was at p<_ 0.05

## Results

Table 1 depicts the demographic data. The mean age of yoga practitioners was 26.4 ± 7.42 years, while that of the sedentary group was 26.8 ± 5.42 years, indicating comparable age distribution between the groups. In terms of gender distribution, females predominated in both groups, accounting for 64.82% of yoga practitioners and 56.81% of sedentary individuals. The majority of participants in both groups were unmarried (85.92% and 86.36%, respectively). Dietary habits revealed that a higher proportion of yoga practitioners (84.42%) followed a vegetarian diet compared to the sedentary group (90.91%), although both groups predominantly adhered to vegetarianism. The prevalence of smoking, alcohol consumption, and drug use was minimal among both groups, with slightly higher rates of smoking (2.27%) and alcohol consumption (4.54%) observed in the sedentary population, whereas drug use (4.02%) was more reported among yoga practitioners. The level of physical activity differed substantially between groups. Among yoga practitioners, 73.37% engaged in regular exercise, 60.80% in walking, 32.66% in jogging, and 12.06% in other activities. Conversely, the sedentary population reported lower engagement in exercise (4.54%), walking (43.18%), jogging (22.72%), and other activities (9.09%). Only 2.01% of yoga practitioners reported regular medication use compared to none in the sedentary group. Previous COVID-19 exposure was noted in 17.09% of yoga practitioners and 11.36% of sedentary individuals.

**Table 1.** presents the demographic characteristics of the study participants, comprising yoga practitioners (N=199) and a non-yoga sedentary population (N=44).

| Sl.No |  | Yoga Practitioner<br>(N=199) | Non-yoga Sedentary population<br>(N=44) |
|---|---|---|---|
| 01 | Age (years) Mean±SD | 26.4 ±7.42 | 26.8±5.42 |
| 02 | Gender<br>Male<br>Female | 35.17<br>64.82 | 43.18<br>56.81 |
| 03 | Marital Status<br>Married<br>Unmarried | 13.56<br>85.92 | 13.63<br>86.36 |
| 04 | Diet<br>Vegetarian<br>Non Vegetarian | 84.42<br>15.57 | 90.91<br>9.09 |
| 05 | Education | Graduate and PG students | Graduate and PG students |
| 05 | Habits<br>Smoking<br>Alcohol<br>Drugs | 1.51<br>1.51<br>4.02 | 2.27<br>4.54<br>0 |
| 06 | Physical activity<br>Exercise<br>Walking<br>Jogging<br>Other | 73.37<br>60.80<br>32.66<br>12.06 | 4.54<br>43.18<br>22.72<br>9.09 |
| 07 | Medication<br>Yes<br>No | 2.01<br>98.49 | 0<br>100 |
| 08 | Covid Exposure<br>Yes<br>No | 17.09<br>82.91 | 11.36<br>88.63 |

All data expressed in percentage (% distribution value). Except for age and education.

Comparative analysis of lung function and cough patterns depicted in Table 2: Yoga practitioners demonstrated significantly superior respiratory performance compared to the sedentary group. Forced Expiratory Volume in one second (FEV1) was higher in yoga practitioners (86.5 ± 10.71) than in non-yoga participants (77.78 ± 20.48), reflecting a 9.56% improvement, which was statistically significant (p ≤ 0.05). Similarly, Forced Vital Capacity (FVC) showed a 10.13% higher value in the yoga group (84.96 ± 9.85) compared to the sedentary population (76.84 ± 19.75), with stronger statistical significance (p ≤ 0.01). The FEV1/FVC ratio, an important indicator of airway function and obstruction, was also significantly higher among yoga practitioners (76.21 ± 7.38) than non-yoga individuals (72.09 ± 14.25), indicating better airway patency and ventilatory efficiency (p ≤ 0.01). The total dry cough count was notably lower in yoga practitioners (7.76 ± 3.16) compared to the sedentary group (6.32 ± 2.98), showing an 18.61% difference that was statistically significant (p ≤ 0.01). In contrast, the difference in wet cough count between the two groups (2.01 ± 1.92 in yoga practitioners vs. 1.89 ± 1.63 in sedentary individuals) was minimal (5.95%) and not statistically significant, suggesting that yoga practice may have a more pronounced effect on reducing irritative or non-productive cough rather than productive cough. Cough duration showed no significant difference between the two groups, with yoga practitioners recording a mean duration of 9.94 ± 0.30 compared to 9.89 ± 0.14 in the sedentary population. This indicates that while yoga practice may influence cough frequency, it does not significantly alter the duration of coughing episodes.

**Table 2.** presents a comparative analysis of lung function parameters and cough patterns measured using the Swaasa App among yoga practitioners (N = 199) and a non-yoga sedentary population (N = 44).

| **Sl.No** | **Variable** | **Yoga Practitioner (N=199)** | **Non-yoga Sedentary population (N=44)** | **Percentage (%) difference** | **Level of Significance** |
|---|---|---|---|---|---|
| 01 | FEV1 | 86.5±10.71 | 77.78± 20.48 | 9.56 | p≤0.05 |
| 02 | FVC | 84.96±9.85 | 76.84± 19.75 | 10.13 | p≤0.01 |
| 03 | FEV1/FVC | 76.21± 7.38 | 72.09± 14.25 | 5.41 | p≤0.01 |
| 04 | Total cough count<br>Dry cough<br>Wet cough | 7.76±3.16<br>2.01±1.92 | 6.32 ±2.98<br>1.89±1.63 | 18.61<br>5.95 | p≤0.01<br>NS |

The data are expressed as mean ± standard deviation, along with percentage differences and levels of statistical significance.NS = Not Significant

Health condition assessment and questionnaire outcomes depicted in Table 3: The overall risk assessment revealed a more favorable respiratory risk profile among yoga practitioners. A higher proportion of yoga practitioners fell into the low-risk category (73.37%) compared to the sedentary group (47.73%), whereas a substantially larger percentage of sedentary individuals were categorized as high risk (52.27%), indicating increased vulnerability to respiratory dysfunction. The majority of yoga practitioners exhibited a normal respiratory pattern (91.46%), while only 68.18% of sedentary participants showed normal patterns. Restrictive and obstructive patterns were markedly lower among yoga practitioners (4.02% and 4.52%, respectively) compared to the sedentary population, where both restrictive and obstructive patterns were reported in 15.91% of participants each. This suggests better pulmonary mechanics and airway integrity in individuals practicing yoga. A large proportion of yoga practitioners fell within the normal severity category (73.87%), whereas only 40.91% of sedentary individuals were classified as normal. High severity respiratory patterns were considerably more prevalent in the sedentary group (20.45%) compared to yoga practitioners (3.02%), indicating more pronounced functional impairment among non-yoga participants. The Lung Health Index showed a mixed distribution; however, yoga practitioners demonstrated a lower proportion in the high-risk category (8.04%) compared to sedentary individuals (31.82%). Although a greater percentage of yoga practitioners were classified under the low Lung Health Index category (59.80%), this may reflect conservative scoring sensitivity within the app, while the sedentary group exhibited a higher clustering in the high-risk zone, suggesting compromised lung health. Yoga practitioners had a lower percentage of individuals categorized as “likely COPD” (26.63%) compared to sedentary participants (36.36%). Similarly, COPD risk stratification revealed that low risk was more prevalent among yoga practitioners (56.78%), whereas a substantially higher proportion of sedentary individuals were classified under high COPD risk (31.82%), highlighting the potential protective role of yoga practice against chronic obstructive airway conditions. Yoga practitioners demonstrated lower asthma likelihood (2.01%) compared to the sedentary population (4.55%). Additionally, asthma risk categorization showed that a majority of yoga practitioners fell into the low-risk category (73.37%), while the sedentary group had a higher proportion of individuals at high asthma risk (27.27%), suggesting increased airway hyperresponsiveness among non-yoga individuals. Analysis of self-reported symptoms indicated a generally lower prevalence of respiratory symptoms among yoga practitioners. Symptoms such as chest pain, frequent cough, wheezing, and difficulty in breathing were more commonly reported in the sedentary population. Notably, wheezing was reported by 15.91% of sedentary individuals compared to 7.04% of yoga practitioners. Exposure to smoke was also higher in the sedentary group (18.18%) compared to yoga practitioners (22.61%), which may contribute to the observed differences in respiratory risk and symptom burden.

**Table 3.**
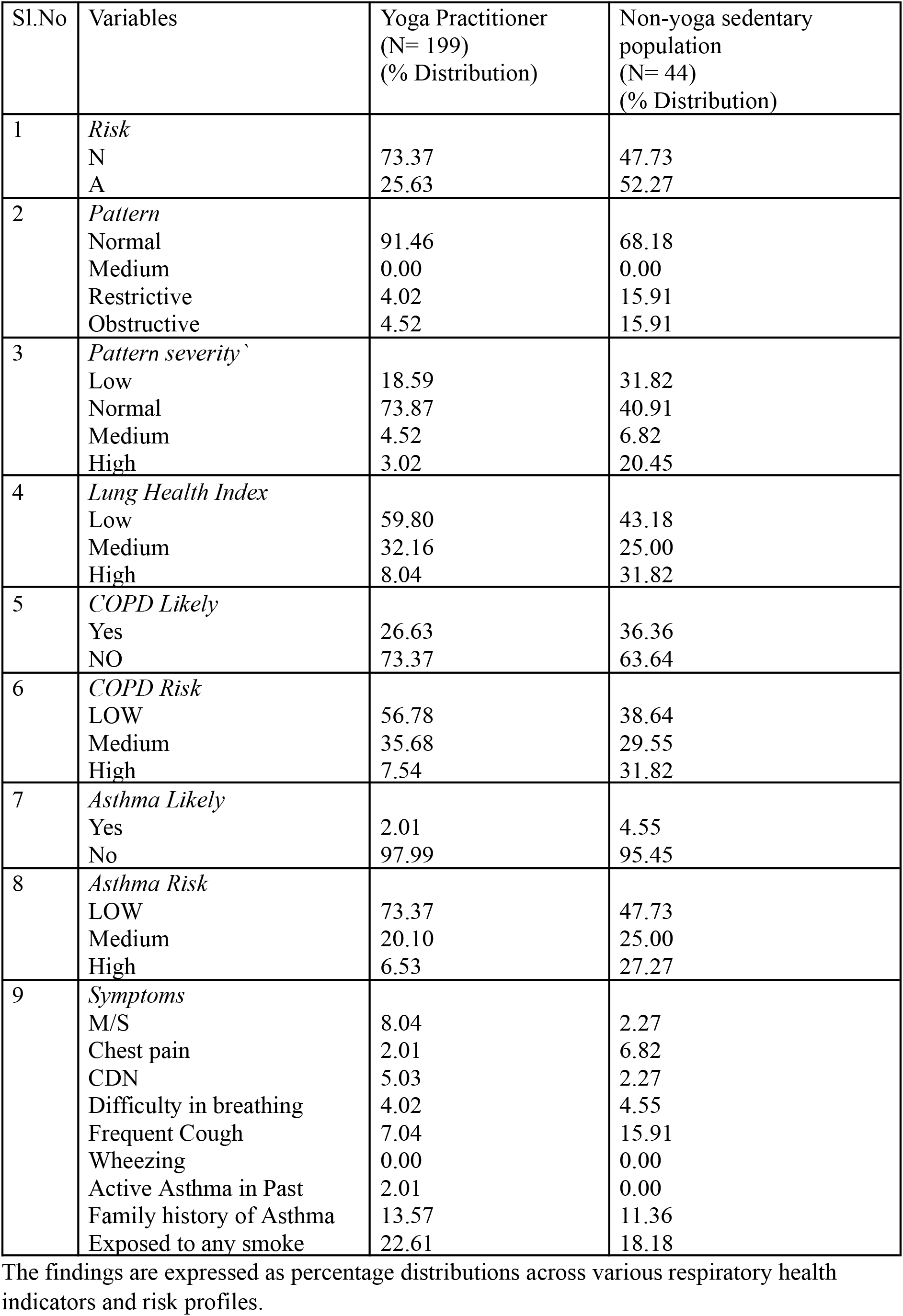
summarizes the health condition assessment and questionnaire outcomes recorded during cough evaluation using the Swaasa App among yoga practitioners (N = 199) and a non-yoga sedentary population (N = 44).

| Sl.No | Variables | Yoga Practitioner<br>(N= 199)<br>(% Distribution) | Non-yoga sedentary population<br>(N= 44)<br>(% Distribution) |
|---|---|---|---|
| 1 | *Risk* |  |  |
|  | N | 73.37 | 47.73 |
|  | A | 25.63 | 52.27 |
| 2 | *Pattern* |  |  |
|  | Normal | 91.46 | 68.18 |
|  | Medium | 0.00 | 0.00 |
|  | Restrictive | 4.02 | 15.91 |
|  | Obstructive | 4.52 | 15.91 |
| 3 | *Pattern severity`* |  |  |
|  | Low | 18.59 | 31.82 |
|  | Normal | 73.87 | 40.91 |
|  | Medium | 4.52 | 6.82 |
|  | High | 3.02 | 20.45 |
| 4 | *Lung Health Index* |  |  |
|  | Low | 59.80 | 43.18 |
|  | Medium | 32.16 | 25.00 |
|  | High | 8.04 | 31.82 |
| 5 | *COPD Likely* |  |  |
|  | Yes | 26.63 | 36.36 |
|  | NO | 73.37 | 63.64 |
| 6 | *COPD Risk* |  |  |
|  | LOW | 56.78 | 38.64 |
|  | Medium | 35.68 | 29.55 |
|  | High | 7.54 | 31.82 |
| 7 | *Asthma Likely* |  |  |
|  | Yes | 2.01 | 4.55 |
|  | No | 97.99 | 95.45 |
| 8 | *Asthma Risk* |  |  |
|  | LOW | 73.37 | 47.73 |
|  | Medium | 20.10 | 25.00 |
|  | High | 6.53 | 27.27 |
| 9 | *Symptoms* |  |  |
|  | M/S | 8.04 | 2.27 |
|  | Chest pain | 2.01 | 6.82 |
|  | CDN | 5.03 | 2.27 |
|  | Difficulty in breathing | 4.02 | 4.55 |
|  | Frequent Cough | 7.04 | 15.91 |
|  | Wheezing | 0.00 | 0.00 |
|  | Active Asthma in Past | 2.01 | 0.00 |
|  | Family history of Asthma | 13.57 | 11.36 |
|  | Exposed to any smoke | 22.61 | 18.18 |

The findings are expressed as percentage distributions across various respiratory health indicators and risk profiles.

## Discussion

The findings of the study highlight that yoga practices are beneficial for respiratory health, or lung health, when compared to a non-yoga sedentary population, as assessed using the Swaasa AI –enabled cough and lung health assessment platform. Also, the study highlights that if an individual is involved in any form of physical activity, then they have a better lung health profile as compared to the ones not involved in any form of physical activity. Collectively, the findings provide technology-assisted evidence supporting the respiratory health benefits of yoga practices.

Yoga practitioners showed higher FEV1, FVC, and FEV1/FVC ratios than the non-yoga practitioners, which shows improved lung volumes and expiratory flow. The findings are also supported by studies done on asana and pranayama improving the respiratory muscle strength, compliance, and ventilatory efficiency. [18-21] Yogic breathing practices also influence parasympathetic modulation and decrease sympathetic overactivity [22,23]

The dry cough count was significantly lower in the yoga practitioner group as compared to the non-yoga practitioners, though the wet cough frequency and cough duration did not differ significantly between the groups. The findings may be because of improved airway hydration, reduced epithelial irritation and better regulation of cough reflex sensitivity through vagal modulation. [24]

The findings also reflect a more favorable risk profile among the yoga practitioner group, reflecting more individuals at low risk and fewer individuals classified in the high-risk category. The findings are supported by previous studies, which show that long-term yoga practice led to improved lung elasticity and thoracic mobility and delayed age-related changes in the respiratory system. [25-27] Also, the findings show normal lung patterns in yoga practitioners, while the restrictive/obstructive patterns were considerably high in the non-yoga practitioner group. Regular practice of asanas leads to improved chest expansion and spinal mobility [28]. Also, self-reported symptoms, including wheezing, frequent cough, chest pain, and dyspnea, were more prevalent in the non-yoga practitioners. Wheezing was reported more than twice as frequently among non-yoga practitioners. These findings are also directly correlated to measures of poorer lung function and higher respiratory risk in the group.

The findings also highlight a preventive or protective effect of yoga, as the yoga practitioner group showed a lower number of individuals categorized as “likely COPD,” as well as lower high-risk stratification for COPD and asthma. The findings are supported by evidence from studies showing that yoga can improve exercise tolerance, reduce dyspnea, and enhance quality of life in individuals with COPD and asthma. [29-31]

### Limitations of the study

An important strength of the study is the use of an AI-driven app, a cost-effective method, to assess the lung health of yoga practitioners and non-yoga practitioners, which enables non-invasive, scalable, and objective assessment of lung health, thus bridging the gap between traditional practices and modern digital technology. However, there are several limitations that the authors acknowledge. The cross-sectional design of the study limits the causal interference and does not establish temporal or dose-response relationships. The unequal sample size may have influenced the statistical power. The small number of samples in non-yoga practitioners may have increased the likelihood of type II error for certain parameters. The detail of the yogic intervention is missing, limiting the dose-intervention interpretation. Also, the study population consisted of a young, relatively healthy adult population with a low prevalence of comorbidities. Hence, the results may not be generalized to older populations, individuals with respiratory disorders, or populations with increased risk exposures.

## Conclusion

The findings of this study demonstrate that yogic practice may be effective in improving lung function parameters such as FVC, FEV1, FEV1/FVC, cough pattern, COPD risk reduction, etc.

## Supporting information

Ethical Approval Letter

## Data Availability

All data produced in the present study are available upon reasonable request to the authors
All data produced in the present work are contained in the manuscript

## Acknowledgements

The authors are grateful to the Director of the Morarji Desai National Institute of Yoga for granting necessary permissions to complete the study. Above all, the authors acknowledge the cooperation and support of their colleagues. The authors are also thankful to the Swaasa team and Salcit Technologies Pvt. Ltd., Hyderabad, for their logistic support for providing the app support. Above all, the cooperation and support of the volunteers is gratefully acknowledged.

