## Supplementary material for "Swaasa an AI-Based Novel Technique to Assess Lung Function in Yoga Practitioners in Comparison to Non-Yoga Practitioners: A Cross-Sectional study": Ethical Approval Letter

मोरारजी देसाई राष्ट्रीय योग संस्थान
MORARJI DESAI NATIONAL INSTITUTE OF YOGA
आयुष मंत्रालय, भारत सरकार
Ministry of Ayush, Govt. of India
68 अशोक रोड, नई दिल्ली- 110001
68, Ashok Road, New Delhi – 110 001

**File No: - MDNIY/YE/MRFM/2020** **Date: - 20/03/24**

To,

Amisha Rawat
M.Sc. Student
Enrolment No- 2152226722
Morarji Desai National Institute of Yoga
68, Ashok Road
New Delhi- 110001

Dear Student,

The Institutional Faculty Cum Research Committee (IFCRC) reviewed and discussed your application to conduct the research work entitled **"Swaasa an AI-Based Novel Technique to Assess Lung Function in Yoga Practitioners in Comparison to Non-Yoga Practitioners : A Cross- Sectional study"** on 15.03.2024.

The following documents were reviewed:

a) The Design of the study and risk involved.
b) Patient Information Sheet and Informed Consent Form.
c) Proposed methods for assessment.

The committee approves the research work to be conducted in its present form.

(Dr. Vandana Singh)
Member Convener, IFCRC

Copy to-

1) Dr. Rameswar Pal, Assistant Professor (H.P.)
2) Ms. Sobika Rao, Assistant Professor (H.A.)
3) Office File
